# Diet pattern determines circulating FGF21 levels while distinct *FGF21* variants influence diet pattern and FGF21 levels

**DOI:** 10.1101/2024.07.08.24309818

**Authors:** Stina Ramne, Mario García-Ureña, Matthew P Gillum, Lars Ängquist, Torben Hansen, Jordi Merino, Niels Grarup

**Author notes:** Correspondence: Niels Grarup.

## Abstract

Experimental and genetic studies suggest that FGF21 modulates macronutrient and alcohol preferences. However, FGF21’s regulation of human appetite remains elusive. To address this gap in translation, we investigated the relationships between plasma FGF21 levels, *FGF21* genetic variation and habitual macronutrient intake in a large human population. We show that the main macronutrient-associated variant rs838133 and the FGF21 cis-pQTL rs838131, both in the *FGF21* gene, are distinct genetic signals. Effect directions also suggest that the influence of *FGF21* variation on macronutrient intake appear more complex than by direct mediation through plasma FGF21. Only when considering this complexity at *FGF21,* is plasma FGF21 estimated to reduce alcohol and increase protein and fat intake using mendelian randomization. Importantly, plasma FGF21 levels are also markedly elevated by high alcohol and low protein intake. This supports the diet-regulatory mechanism of FGF21 in humans, but highlights the need for mechanistic characterization of the *FGF21* genetic region.

## Introduction

Diet is one of the main modifiable risk factors for cardiometabolic diseases. A better understanding of the mechanisms underlying dietary behaviors is critical to identify strategies to prevent and manage these diseases. A promising biological regulator of dietary preferences is the hepatic hormone fibroblast growth factor 21 (FGF21). FGF21 exerts a wide range of beneficial metabolic effects (1), and although the development of FGF21-based drugs for treatment of mainly metabolic liver disease is advancing (2), a lot of questions regarding FGF21’s endocrine regulation of dietary preferences in humans remain unanswered.

Experimental studies have shown that administration or overexpression of FGF21 in animals reduce alcohol and sugar preference while promoting protein feeding (3–6). Similar effects have been suggested in a human trial, where FGF21 receptor activation reduced sweet taste preference (7). On the other hand, alcohol and sugar intake and protein restriction, induce a robust FGF21 response in humans (8–13). This points to a negative feedback regulation of appetite and intake of these macronutrients, acting to protect us from an imbalanced consumption (14, 15).

Most research of this diet-regulatory mechanism comes from animal models, but human relevance of such a mechanism is supported by the fact that the *FGF21* locus is the strongest signal in genome- wide association studies (GWASs) of macronutrient intake (16–20). Nevertheless, there are gaps in the translation of this research that potentially could be addressed by incorporating individual level data on plasma FGF21 protein levels, genetic variation and habitual dietary intake in large population cohorts. For example, experimental work suggests that FGF21’s control over sweet taste preference is secondary to its control over protein preference (6, 12, 21), but this has not been studied in the context of the complex human diet. Furthermore, although there is a clear link between FGF21 and alcohol intake (5), the experimental research on FGF21’s role in macronutrient appetite generally neglects alcohol, which sometimes is referred to as the fourth macronutrient. The human literature on plasma FGF21 levels related to dietary intake in humans is also generally limited to the acute effects of small dietary experiments, whereas the research on long-term self-selected complex dietary habits is limited to studies of *FGF21* genetic variation. Thus, the association between habitual dietary intake and plasma FGF21 levels has not been studied, which is important as diet may alter FGF21 levels, and FGF21 levels may alter diet.

In addition, there appears to be a discordance in direction of effects in previous observations of genetic *FGF21* variation and macronutrient intake. The lead diet-associated *FGF21* rs838133 A-allele associates with higher relative carbohydrate and sugar intake but lower protein and fat intake (16, 18–20). This allele has in functional studies been demonstrated to cause elevated plasma FGF21 levels (22). The same was shown for rs838145 (17). Given that FGF21 administrations in animals shift preferences from sugar to protein (6, 21), we would expect FGF21-increasing alleles to associate with macronutrient intake in the opposite direction, as the most intuitive would be that a genetic variant influences a behavior through altered protein levels. This discordant directionality of *FGF21* variants is an intriguing conundrum, that may have implications for our ability to estimate the casual effects of FGF21 on macronutrient preference in humans using mendelian randomization (MR).

By mapping out the relationships between plasma FGF21 levels, *FGF21* genetic variation and habitual macronutrient intake in an observational setting, we aimed to investigate the directionality of *FGF21* genetic associations with macronutrient intake, whether the postprandial relationship between macronutrient intake and plasma FGF21 is preserved in the long-term perspective, the macronutrient priorities of FGF21’s dietary regulation, and lastly the possibilities for genetically estimating the casual effect of FGF21 on macronutrient preferences in humans.

## Results

### Evidence of distinct *FGF21* signals influencing macronutrient intake and plasma FGF21 levels

First, we aimed to better understand previous observations at the *FGF21* genetic region. We used summary statistics from GWASs of plasma FGF21 levels (23) and macronutrient intake (20) to investigate the confluence between genetic variants influencing these phenotypes. Visual inspection at the *FGF21* region suggested distinct genetic signals for plasma FGF21 levels and macronutrient intake (Figure 1), despite that the two lead variants, the intronic rs838131 variant (MAF 49%) for plasma FGF21 levels and the synonymous rs838133 variant (MAF 43%) for macronutrient intake, are located only 1,148 bases from each other (LD R^2^=0.33, D’=0.62). Fine-mapping coupled with functional annotations to find probable causal variants showed no overlap between the identified variants for plasma FGF21 levels and macronutrient intake (Supplemental table 1, Supplemental figure 1). Colocalization analyses of plasma FGF21 levels and macronutrient intake in this genetic region demonstrated a probability of >0.99 that there is an association with both studied traits explained by two independent variants and minimal probability that these traits are colocalized (Supplemental table 2). Taken together, these investigations of GWAS summary statistics provide evidence that there are two distinct genetic signals at the *FGF21* locus for plasma FGF21 levels and macronutrient intake.

**Figure 1.**
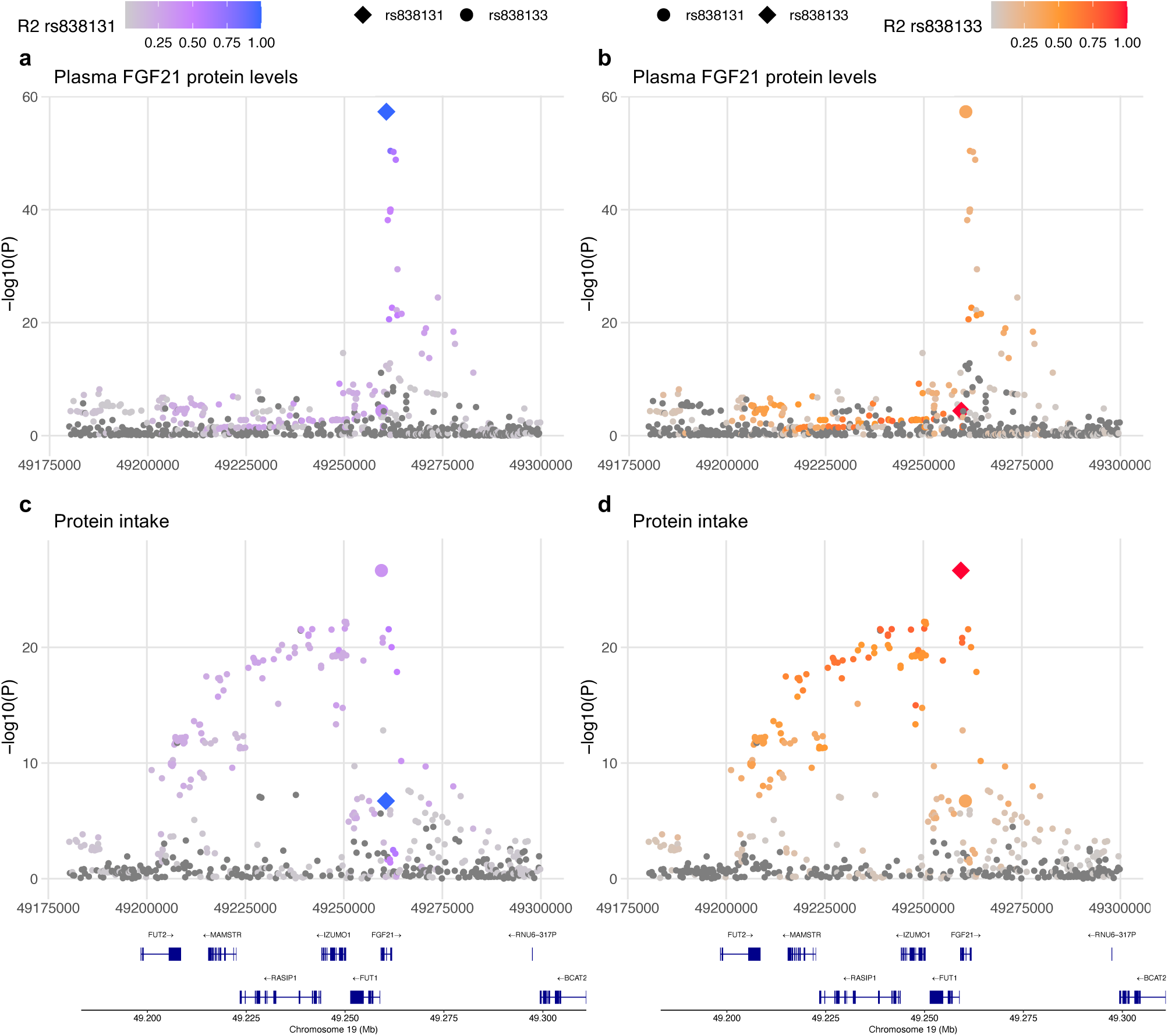
Regional plots of the genetic associations at the FGF21 locus. Plots display position 49180000 to 49300000 at chromosome 19. **a**, Associations with plasma FGF21 with color-scale representing LD R^2^ with rs838131. **b**, Associations with plasma FGF21 with color-scale representing LD R^2^ with rs838133. **c**, Associations with protein intake with color-scale representing LD R^2^ with rs838131. **d**, Associations with protein intake with color-scale representing LD R^2^ with rs838133.

### Phenotypic variation related to the distinct *FGF21* variants

Next, we compared the two *FGF21* variants’ associations with plasma FGF21 levels and macronutrient and alcohol intake data measured with repeated 24-hour recalls (24HRs) expressed as percentages or total energy intake (E%) in the UK Biobank (UKBB). As expected, the rs838131 variant was much more strongly associated with higher plasma FGF21 levels than the rs838133 variant, and this was minimally altered when conditioning on rs838133 (Figure 2a-c). Similarly, the rs838133 variant was much more strongly associated with macronutrient intake than the rs838131 variant, predominantly with low protein and fat intakes and high free sugar and alcohol intake, and these associations were minimally altered when conditioning on the rs838131 variant (Figure 2d-f, Supplemental figure 2). These analyses strengthen the notion of two distinct genetic signals, where rs838131 alters plasma FGF21 levels and rs838133 alters macronutrient intake.

**Figure 2.**
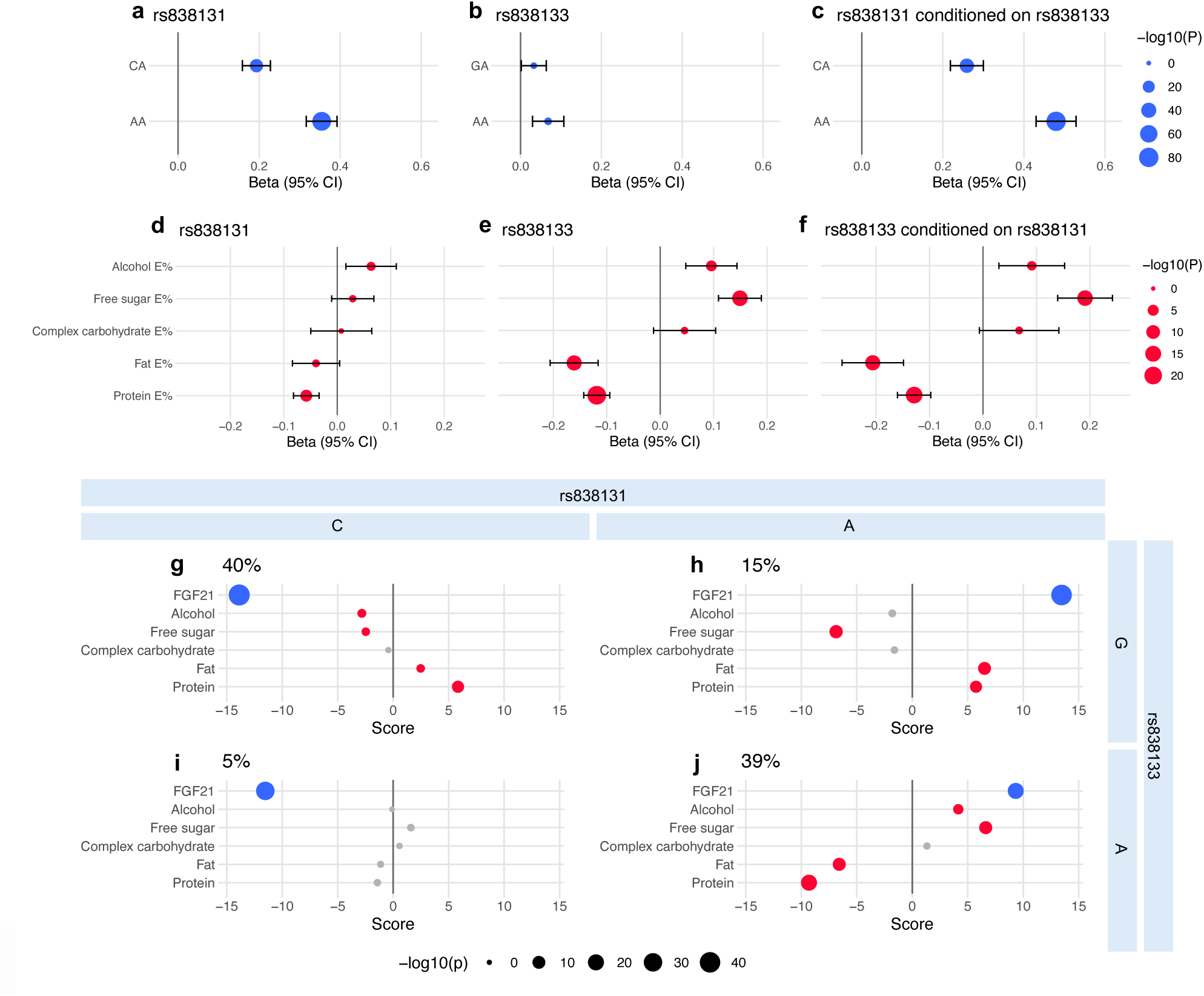
Association of the rs838131 and rs838133 variants in association with plasma FGF21 levels and macronutrient intake. Associations between **a**, rs838131, **b**, rs838133, and **c**, rs838131 conditioned on rs838133 and plasma FGF21 levels. Analyses with plasma FGF21 levels (a-c) are adjusted for age, sex, study center, fasting time, BMI and 10 genetic PCs (n=38,779). Associations between **d**, rs838131, **e**, rs838133, and **f**, rs838133 conditioned on rs838131 and macronutrient intake. Analyses with macronutrient intake (d-f) are adjusted for age, sex, study center, BMI, total energy intake and 10 genetic PCs (n=146,241-157,508). Haplotype score test of plasma FGF21 levels and macronutrient intake levels with haplotypes generated from the two FGF21 variants (g-j). G) CG haplotype, H) AG haplotype, I) CA haplotype, J) AA haplotype. Blue dots represent associations with plasma FGF21 levels, adjusted for age, sex, study center, fasting time, BMI and 10 genetic PCs. Red dots represent associations with macronutrient intake, adjusted for age, sex, study center, total energy intake, BMI and 10 genetic PCs. Grey dots are non-significant score tests.

We estimated haplotypes of the two *FGF21* variants and performed score tests with plasma FGF21 levels and macronutrient intakes (Figure 2g-j). The most common haplotype, rs838131-C and rs838133-G (CG, frequency 40%) was strongly associated with lower FGF21 plasma levels and associated with lower alcohol and free sugar intake, and higher protein and fat intake. In contrast, the rs838131-A and rs838133-G haplotype (AG, frequency 15%) showed an opposite association with plasma FGF21 levels, while the associations with macronutrient intake remained in the same direction as the CG haplotype. The common rs838131-A and rs838133-A haplotype (AA, frequency 39%) was associated with higher plasma FGF21 levels, but to a reduced degree than the AG haplotype, while this haplotype most strongly associated with macronutrient intake, positively with alcohol and free sugar and negatively with protein and fat. The rs838133-A and rs838131-C haplotype (CA, frequency 5%) associated negatively with plasma FGF21 levels but not with macronutrient intake. These haplotype analyses suggest that the effect of the rs838131 A allele on plasma FGF21 levels is attenuated by the rs838133 A allele, whereas the effect of the rs838133 A allele on macronutrient intake is strengthened by the rs838131 A allele. The same conclusions were reached from comparison of all nine possible genotype group combinations of the rs838131 and rs838133 variants (Supplemental figure 3).

### Phenotypic associations between plasma FGF21 and macronutrient intake

The differentiation between the *FGF21* genetic signals for plasma FGF21 levels and macronutrient intake, respectively, raises the question whether an association between habitual macronutrient intake and plasma FGF21 levels exists, or if this only is reflected in the acute response to intake of specific macronutrients. We investigated the cross-sectional associations between macronutrient intake and plasma FGF21 levels in the UKBB. We found that a higher intake of alcohol and free sugar, while lower intake of protein, complex carbohydrate and fat, were associated with higher plasma FGF21 levels (Figure 3a). We also studied intake of macronutrients modeled as substitutions (24) to come closer at understanding the macronutrient priorities in FGF21 regulation. These analyses demonstrated that both lower protein intake and higher alcohol intake, in substitution for any other macronutrient, were strongly associated with higher plasma FGF21 levels (Figure 3b). Higher alcohol intake at the expense of protein intake was most strongly associated with high plasma FGF21 levels. The general direction of these associations aligns with the effects that experimental macronutrient manipulations have on postprandial FGF21 responses, indicating that plasma FGF21 levels cross-sectionally reflect a consequence of macronutrient intake.

**Figure 3.**
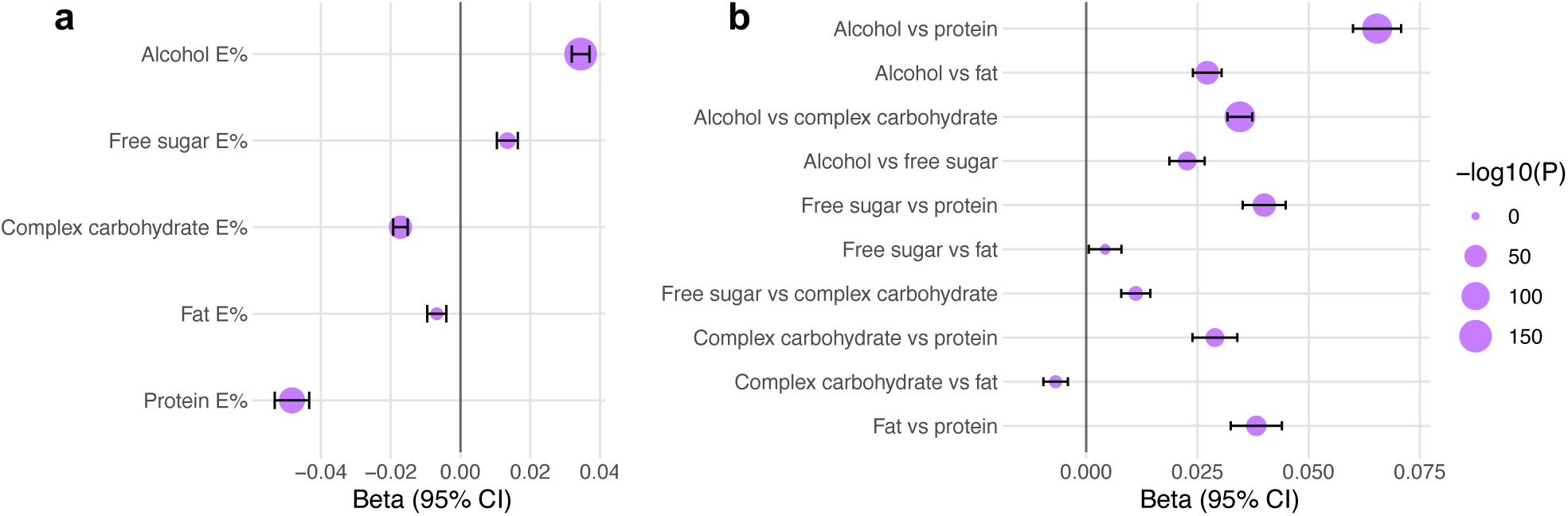
Associations between macronutrient intake and plasma FGF21 levels. **a**, Association for intake of isolated macronutrients and **b**, associations for macronutrient intake modeled as pairwise substitutions. Analyses were adjusted for age, sex, study center, fasting time, BMI and total energy intake in addition to macronutrients for substitution modelling (n=21,604).

We further showed that the effect sizes of the associations between macronutrient intake and plasma FGF21 were somewhat attenuated but still highly significant when restricting to individuals with obesity (BMI ≥30) (Supplemental table 3) and when restricting to individuals that had fasted for at least 6 h before given their blood sample (Supplemental table 4). Furthermore, these associations were not substantially affected by adjustment for either of the distinct *FGF21* variants rs838131 and rs838133, nor if adjusting for a weighted SNP-score based on all seven protein quantitative traits loci (pQTLs) identified for plasma FGF21 levels in the UKBB (23) (Supplemental table 5). This indicates that macronutrient intake is a stronger determinant of plasma FGF21 levels than *FGF21* genetic variation.

### Estimates of FGF21’s effects on macronutrient intake using mendelian randomization

The direction of the associations with the *FGF21* variants appears to not fit with a potential mechanism where *FGF21* variants act through plasma FGF21 levels to alter macronutrient intake given previous experimental evidence, as further supported by the lack of colocalization between these traits. This potentially impedes estimation of the casual effects of FGF21 on macronutrient intake using MR with these variants as instrumental variables (IVs) (25). To investigate this, we first performed one-sample two-stage least-squares MR estimating the potentially causal effect of FGF21 on macronutrient intake substitutions in the UKBB. We compared different IVs: the single plasma FGF21 cis-pQTL (rs838131), a weighted SNP score of 7 FGF21 pQTLs (23), and a weighted SNP score of only 6 FGF21 trans-pQTLs, thus excluding rs838131. We did not study rs838133 as an IV, as it explains little variation in plasma FGF21 levels and rather is associated with the outcome. As expected, using the rs838131 as IV, the MR-estimates suggested that FGF21 increases alcohol and free sugar intake at the expense of fat intake (Figure 4a). When using the weighted SNP score, plasma FGF21 was instead estimated to reduce alcohol intake in replacement of protein, which was further strengthened when using only the FGF21 trans-pQTLs SNP score (Figure 4b-c). This model of trans-pQTLs demonstrated MR-estimates of FGF21 reducing alcohol in substitution for any macronutrient, as well as increasing protein intake at the expense of any other macronutrient (Figure 4c), which align with the effects on diet preferences from experimental FGF21 research. We show similar effects of plasma FGF21 on macronutrient intake without modelling substitutions (Supplemental figure 4).

**Figure 4.**
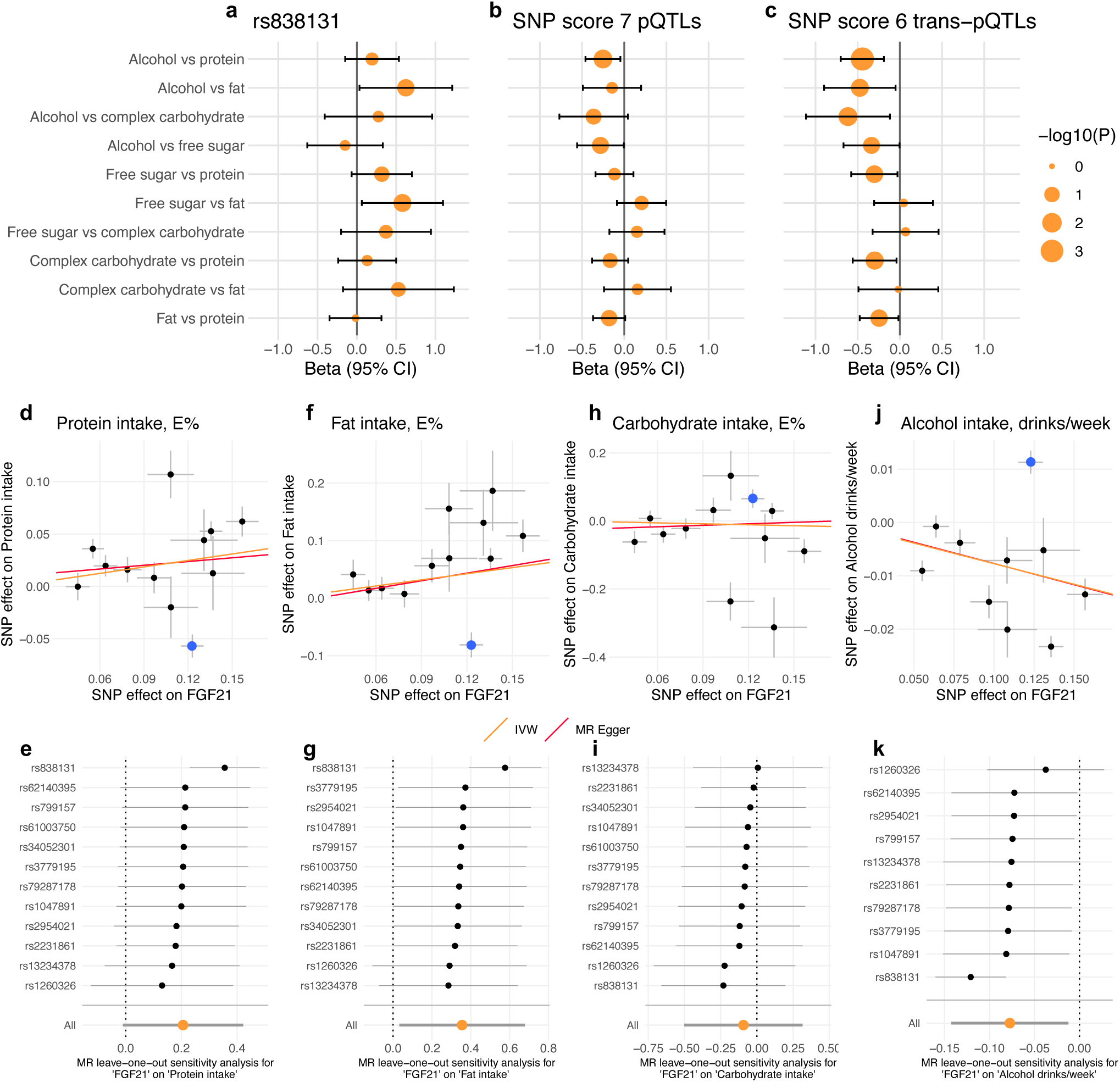
MR analyzes estimating the effects of plasma FGF21 on macronutrient intake. **a**-**c** demonstrates one-sample MR analyses estimating the effect of plasma FGF21 on macronutrient intake substitutions using different IVs: **a**, the FGF21 cis-pQTL (rs838131), **b**, all 7 FGF21 pQTLS composed into a weighted score, and **c**, only the 6 trans-pQTLs (excluding rs838131) composed into a weighted score. Analyses were adjusted for age, sex, study center, fasting time, BMI, total energy intake and 10 genetic PCs in addition to macronutrients for substitution modelling (n=15,512- 16,400). **d**-**k** demonstrates two-sample MR analyses estimating the effect of plasma FGF21 on macronutrient intake using GWAS summary statistics. Scatter plots depicting the relationship between each variant’s effect on both plasma FGF21 and **d**, protein intake, **f**, fat intake, **h**, carbohydrate intake and **j**, alcohol intake (the rs838131 variant is highlighted in blue). Sensitivity leave-one-out analyses are presented for **e**, protein intake, **g**, fat intake **i**, carbohydrate intake, and **k**, alcohol intake, wherein MR effect estimates represent those when excluding the indicated variant as IV from the model.

These one-sample MR results were replicated in two-sample MR analyses using GWAS summary statistics of plasma FGF21 levels (23), protein, fat and carbohydrate intake (20), as well as alcohol use in drinks per week (26). We performed leave-one-out sensitivity analysis and observed that only when excluding rs838131 as IV, could we find MR support for that plasma FGF21 levels increase protein and fat intake and reduce alcohol intake (Figure 4d-k, Supplemental figure 5), aligning with findings in experimental animal models. We found no effect of FGF21 on carbohydrate intake in two-sample MR analyses. MR heterogeneity was consistently reduced when excluding the rs838131 variant as IV (Supplemental table 6). All studied IVs, including rs838131, passed the Steiger filtering test indicating that they influence the outcome through the exposure and not the other way around.

## Discussion

This study maps the relationship between genetic variation, plasma FGF21 levels and self-reported habitual macronutrient intake in a large free-living population. Combining these data has only recently become possible owing to new large-scale proteomic resources. We find distinct genetic signals at the *FGF21* locus for plasma FGF21 levels and macronutrient intake. We also demonstrate bidirectional effects between macronutrient intake and plasma FGF21 levels similar to experimental evidence, where plasma FGF21 are markedly elevated by primarily high alcohol and low protein intake, whereas MR analyses supports an effect of plasma FGF21 on reducing alcohol intake and increasing protein and fat intake.

Two different genetic loci in the *FGF21* gene are responsible for two separate effects. One led by rs838131, responsible for increasing FGF21 levels. The other led by rs838133, responsible for altering macronutrient preferences. The intronic rs838131 variant is until now unreported in PubMed or the GWAS catalog. Thus, we have limited knowledge of its molecular consequences. The synonymous rs838133 variant, however, has been shown to yield higher FGF21 plasma levels owing to faster translation and increased protein stability (22). Despite this, we show that this variant is only weakly associated with plasma FGF21 levels.

This study also demonstrates that *FGF21* genetic variants associate with plasma FGF21 levels and macronutrient intake in opposite direction to what would be the most intuitive pathway, i.e. that a genetic variant acts through altered protein levels to influence a behavior. However, the observed directions fit with a scenario where *FGF21* variation alters macronutrient intake, and subsequent effects on plasma FGF21 levels of such intake, but this is less intuitive for a cis-gene-protein relationship. Other mechanisms explaining how *FGF21* genetic variation influences macronutrient preference could be at play, for example the case of FGF21 resistance. This has mainly been discussed in the context of obesity (27), but Bayoumi et al. have postulated it in relation to rs838133 (22). If we similarly speculate that chronically elevated FGF21 levels owing to *FGF21* genetic variation could result in FGF21 resistance in the brain, then the FGF21 signal to shift dietary preferences may be attenuated. This could fit with the observed direction of effect of the *FGF21* variants in this study, but if this was the case we would expect to observe colocalization of the genetic signals for plasma and macronutrient intake. Alternatively, as the Olink measurements of plasma FGF21 levels in the UKBB most likely reflect total FGF21, including the inactive form (28), it is possible that the associations for the *FGF21* variants are explained by high levels of inactive FGF21. Future genetic investigations of both total and active circulating levels of FGF21 would be valuable and molecular characterization of the *FGF21* variants is needed to determine how they affect diet preferences. This is important to understand if continued exposure to high FGF21 levels can lead to FGF21 resistance, since this may be crucial for the implementation of FGF21-based therapies.

The current findings also raise concerns for using MR to study the potentially causal effects of FGF21. It is generally preferred to use cis-pQTLs as IVs when studying causal effect of proteins, as this suggests a more certain relationship between the IV and exposure (29). Although, evidence of colocalization should preferably also be present (30), which is not the case of the FGF21 cis-pQTL rs838131. In both one- and two sample MR, only when excluding the rs838131 variant as IV, could we demonstrate MR-estimated effects of FGF21 on reducing alcohol intake and increasing protein and fat intake. This direction of effect aligns with experimental evidence and suggests that FGF21 might have similar effects on macronutrient preference in humans as seen in animals. In contrast, MR using solely rs838131 as IV, generated weak estimates of FGF21 in the opposite direction. A previous two-sample MR study using another single *FGF21* variant as IV, identified as the lead FGF21 cis-pQTL in a Swedish population (rs2548957; LD with rs838131 r^2^=0.63, D’=0.94), also found estimates of FGF21 increasing alcohol intake (31, 32). These MR estimates using solely the *FGF21* locus oppose experimental data and the MR estimates from genome-wide IV selection, and are thus likely not a correct representation of causal effects. Nevertheless, it should be noted that several of the other IVs in our MR analyses may not be unproblematic. Variants in both *GCKR* and *MLXIPL* have previously been identified in macronutrient intake GWASs too (20), and their gene products are in various ways involved in carbohydrate metabolism (33, 34). The product of *MLXIPL,* carbohydrate response element binding protein (ChREBP), is also necessary for generating the FGF21 plasma response following intake of both sugar (3) and alcohol (35). Thus, the MR analyses should be interpreted with caution, even though they are supported by experimental evidence, and underlines the need for rigorous consideration of mechanisms and assumptions for IV selection.

While our MR analyses suggest that there is an effect of FGF21 on macronutrient intake, we also demonstrate that the cross-sectional relationship between macronutrient intake and plasma FGF21 levels, primarily reflects the effect that macronutrient intake has on FGF21 levels. These strong associations indicate that long-term habitual diet has similar effects on fasting plasma FGF21 levels as how a macronutrient load affects the acute postprandial FGF21 response. This interpretation is supported by the fact that macronutrient intake was assessed with multiple 24HRs spread out months apart, not in direct connection to the plasma samples, and that these associations remained when restricting to blood samples taken after minimum 6 h of fasting. Long-term effects of dietary habits on fasting FGF21 has previously been suggested from a few randomized dietary interventions in humans (11, 36, 37), but distinguishing between randomly allocated diet interventions and self-selected diet is of importance in this context, as FGF21 also regulates macronutrient selection. Ultimately, we show that habitual macronutrient intake associates with plasma FGF21 levels independent of *FGF21* genetic variation, highlighting the strength of which macronutrient determines plasma FGF21 levels.

Lastly, this study increases our understanding of the prioritized nutrients under FGF21’s control. Through the use of macronutrient substitution models we were able to take into consideration that intake data is compositional, i.e. that increasing intake of one nutrient inherently displaces another (38). This is of particular importance when studying FGF21, as several macronutrients are involved in its regulation, primarily alcohol, sugar and protein. With substitution modeling we show that free sugar intake was only strongly associated with plasma FGF21 when substituting protein, and that higher alcohol intake was strongly associated with higher FGF21 levels even in substitution for free sugar, together suggesting that free sugar intake is less deterministic of plasma FGF21 levels than alcohol and protein intake. This is in line with our previous meal study testing macronutrient interactions on postprandial FGF21 secretion in humans, where we found that protein outweighed sucrose in the secretory regulation of FGF21 (12). Mice experiments have also indicated that the effect of FGF21 on sugar intake is secondary to its effects on protein intake (6, 21). Alcohol has been neglected in these previous macronutrient comparisons, but as alcohol is the strongest acute inducer of acute FGF21 secretion (9, 39), it is not surprising to find the highest plasma FGF21 levels among the macronutrient substitutions defined by high alcohol intake. We also adapted the method for modeling macronutrient substitutions as outcomes in genetic analyses. Here, the associations between the diet-associated rs838133 variant and macronutrient substitutions were contrastingly stronger for free sugar and complex carbohydrate than alcohol in one direction, while both protein and fat were strongly associated in the opposite direction. On the other hand, with MR using IVs from across the genome but excluding rs838131, we found that FGF21 mainly reduces alcohol intake and increases protein and fat intake, with no effect on carbohydrate intake. Taken together, genetic variation at the *FGF21* gene specifically appears mainly associated with an imbalance in carbohydrate and protein intake, while FGF21 variation in entirety (plasma level measurements or IV estimation from across the genome) appears more strongly associated with alcohol and protein imbalance. These divergent observations of macronutrient associations with plasma FGF21 and the *FGF21* locus are not yet understood and requires further investigation.

In conclusion, this study provides evidence of two independent *FGF21* signals accounting for elevating plasma FGF21 levels and altering macronutrient preferences. Effect directions does not suggest that the influence of genetic *FGF21* variation on macronutrient intake are directly mediated by FGF21 levels, for yet unknown reasons. Using MR, we demonstrate support for that FGF21 reduces alcohol and increases protein and fat intake in humans, alike observations in animal models. Furthermore, plasma FGF21 levels are markedly elevated by higher alcohol and lower protein intake, indicating that FGF21 production is regulated by the dietary behaviors that FGF21 is postulated to regulate. These findings are of great significance for the precision nutrition field, as genotype-based targeted approaches based on relevant *FGF21* variation, may have the potential to increase the success rate of dietary interventions to prevent and manage cardiometabolic disorders.

## Method

### UKBB study population

The UKBB is a prospective cohort study covering 502,166 individuals aged 40-69 collected between 2006 and 2010 from 22 different sites in the UK. All participants signed informed consent prior to participation. Participants took part in a wide range of assessments including sociodemographic questionnaires, lifestyle questionnaires, anthropometrics, and blood samples, etc. (40).

### UKBB genotype data

Genotyping of the UKBB was mainly performed using the Applied Biosystems UK Biobank Axiom Array, but also in part the Applied Biosystems UK BiLEVE Axiom Array by Affymetrix. Quality control, imputation and other details are provided in the UKBB method documentation. In the genetic analyses of this study, we only included individuals that were of British ancestry (41). We aligned the two studied *FGF21* genetic variants, rs838131 and rs831833, so that we assessed the effect of the FGF21-increasing alleles. We calculated weighted SNP-scores of the pQTLs for FGF21 identified in Sun et al (23), by weighting the genotype data of each SNP by their effect size in the discovery sample and summed together. We calculated one weighted SNP-score with all seven pQTLs and one SNP-score with only the trans-pQTLs, excluding the cis-pQTL rs838131.

### UKBB plasma FGF21 protein measurements

Measurement of plasma FGF21 protein levels were performed using the Olink platform as part of the UKBB-PPP initiative, wherein 2,922 plasma proteins were measured in 53,058 participants of the UKBB (23). Olink utilizes the proximity extension assay technology for targeted proteomics analysis. Protein abundances are expressed as normalized protein expression (NPX) levels. More details are provided in the UKBB method documentation. In this study, we only utilized FGF21 measurements from instance 0, i.e. at the initial assessment visit. Participants were not required to fast before this blood collection, and the mean (SD) fasting time at sample collection was 3.8 (2.5) h. Therefore, all analyses of plasma FGF21 protein levels are adjusted for fasting time.

### UKBB diet assessment

Dietary intakes in the UKBB were assessed using the web-based 24HR method Oxford WebQ, which asks about intake of 238 different food and drink items consumed within the last 24 hours. At least one 24HR assessment of intake has been provided from 210,856 UKBB participants, while most have provided more than one assessment, up to a maximum of five assessments (42). Invitations to fill out web-based 24HRs after attending the baseline assessment were sent out with a gap of 3-4 months between each to capture intake across the seasons. Food and drink intake frequencies were multiplied by standard portion sizes and the nutrient composition of each item to translate the data into daily nutrient intakes (42). In this study, we excluded 24HR assessments with reported energy intakes above 20,000 kJ for men and 18,000 kJ for women (42). We calculated the average nutrient intake across each individual’s varying number of 24HR assessments for total energy intake and the macronutrients of interest: total fat, total protein, total carbohydrate, free sugar (defined as added sugar plus naturally occurring sugar in fruit juices, honey and syrup), and alcohol. We further calculated percentages of energy intake of these macronutrients and calculated intake of *complex carbohydrate* by subtracting intake of free sugar from intake of total carbohydrate. Thus, we studied the E% of fat, protein, complex carbohydrate, free sugar and alcohol, which together makes up 100% of the total energy intake.

### UKBB statistical analyses

Linear regression analyses between macronutrient intake E% and plasma FGF21 levels were adjusted for the following covariates: age, sex, study center, fasting time, BMI, and total energy intake. We also modelled the associations between macronutrient substitutions and plasma FGF21 levels using the leave-one-out method as described by Song et al (24). For example, to model higher intake of alcohol in substitution for protein, the associations between alcohol intake and plasma FGF21 levels were in addition to the covariates, adjusted for intake of fat, complex carbohydrate and free sugar, leaving out protein from the model (24). Thus, the effect of alcohol on FGF21 is only allowed to vary at the expense of protein intake, as all other macronutrients are held constant. This substitution modelling was performed for each possible pairwise macronutrient substitution. These analyses were also performed stratified for BMI <30 kg/m^2^ and ≥30 kg/m^2^, as well as stratified for fasting time <6 h and ≥6 h. We also evaluated the influence of adjusting for the *FGF21* genetic variants on the associations between macronutrient substitutions and plasma FGF21 levels.

When analyzing the association between the *FGF21* genetic variants and plasma FGF21 levels the following covariates were included in the model: age, sex, study center, fasting time, BMI, and 10 genetic principal components (PCs). We used an additive genetic model and the *FGF21* genetic variants were aligned to the FGF21-increasing allele, which for both the rs838131 and rs838133 variants were the A allele.

The associations between the *FGF21* genetic variants and macronutrient intake E% were adjusted for the following covariates: age, sex, study center, BMI, total energy intake, and 10 genetic PCs. We also adapted the leave-one-out method to study macronutrient substitutions as outcomes when investigating the associations between genetic variants and macronutrient intake. If modeling higher alcohol intake in substitution for protein, the associations between a genetic variant and alcohol intake were, in addition to the listed covariates, adjusted for intake of fat, complex carbohydrate, and free sugar – leaving out protein. With this approach we add the macronutrients as dependent variables alongside the genetic variant. This can be interpreted as if the both the exposure (genetic variants) and the outcome (e.g. alcohol intake) are adjusted for these macronutrient covariates, and thus, represent the alcohol-protein substitution. Throughout the study we present associations with macronutrients modelled both in isolation and as substitutions, to demonstrate that the results and conclusions are robust to the interpretational differences between these two models.

We used the R-package *haplo.stats* to estimate haplotypes of the two *FGF21* variants s838131 and rs838133, and test their respective association with plasma FGF21 levels and macronutrient intakes. We used *haplo.score* tests (43) adjusted for age, sex, study center, fasting time, BMI, and 10 genetic PCs for plasma FGF21 levels, and adjusted for age, sex, study center, BMI, total energy intake, and 10 genetic PCs for macronutrient intakes.

We also constructed genotype groups of all nine possible combinations of the rs838131 variant (CC, AC or AA) and the rs838133 variant (GG, AG, AA). We investigated how these groups associated with plasma FGF21 levels and macronutrient intake in comparison to the reference group rs838131 CC and rs838133 GG. Associations with plasma FGF21 were adjusted for age, sex, study center, fasting time, BMI, and 10 genetic PCs, and associations with macronutrients were adjusted for age, sex, study center, BMI, total energy intake, and 10 genetic PCs.

We used the *IVreg* R-package to conduct one-sample two-stage least-squares MR analyses estimating the causal effect of plasma FGF21 on macronutrient intake (modeled both isolated and as substitutions). A variety of IVs were compared: the FGF21-associated rs838131 variant (the cis pQTL), the total pQTL weighted SNP-score, and the trans-pQTL SNP-score excluding the rs838131 variant. The MR analyses were adjusted for age, sex, study center, fasting time, BMI, total energy intake, and 10 genetic PCs.

All analyses have been performed in R version 4.3.3.

### Analyses of GWAS summary statistics

We utilized summary statistics data of common genetic variants from the plasma FGF21 GWAS within the UKBB-PPP project by Sun et al. (23) and GWAS summary statistics on protein, carbohydrate and fat intake by Merino et al. (20) to plot the region covering both the entire macronutrient signal and the plasma FGF21 signal on chromosome 19 (chr19:49180000 to chr19:49300000, GRCh37) together with LDproxy data from the European 1000 Genomes panel for each respective lead variant.

This region was then fine-mapped using the R-package *CARMA* (44) for plasma FGF21, protein intake, fat intake and carbohydrate intake. We chose this fine-mapping method since it can correct for LD differences between summary statistics from meta-analyses and an external reference panel. Here we ran CARMA with the default settings utilizing the European 1000 Genomes reference panel Phase 3 version 5 and its functional annotations. We only deemed as probable causal variants those that were genome-wide significant (P<5×10^-8^) and had posterior inclusion probabilities (PIPs) >0.1. Variants with a PIP >0.5 were plotted in the supplemental regional plots.

We ran colocalization analyses under the single causal variant assumption with the *coloc* R-package (45) to test for colocalization between plasma FGF21 levels and protein, fat and carbohydrate intake in the genomic region previously described. In addition to using the summary statistics from the Sun et al. FGF21 GWAS (23) and the macronutrient GWASs by Merino et al. (20), we ran the same colocalization analysis with summary statistics from the earlier Merino et al. GWASs on protein intake, fat intake and carbohydrate intake in the CHARGE consortium (18). This to test for colocalization with no overlapping individuals between the macronutrient traits and the plasma FGF21 levels, i.e. in two independent populations. The colocalization analyses were run using the approximate Bayesian factor (abf) method using the default prior probabilities (45).

We used the R-package *TwoSampleMR* to run two-sample MR analyses (46) using GWAS summary statistics, estimating the effect of plasma FGF21 levels (23) on protein, fat and carbohydrate intake (20), as well as alcohol use in drinks per week (26). We identified IVs for plasma FGF21 levels as genome-wide significant variants (P<5×10^8^) and with LD clumping using the European 1000 Genomes reference panel (distance 500 kb, R^2^ threshold 0.01). Two-sample MR were performed using both the inverse variance weighted method and the Egger method. We evaluated each IV’s influence on these MR-estimates by testing every IV individually and using a leave-one-out approach. We also evaluated the heterogeneity of each tested MR model, as well as the directionality of each IV using the Steiger filtering function.

## Data availability

The UKBB data is made publicly available after an application procedure as described here http://www.ukbiobank.ac.uk/using-the-resource/.

GWAS summary statistics of FGF21 plasma protein UKBB-PPP project are available at https://www.synapse.org/Synapse:syn51365301.

Summary statistics of meta-analyzed GWASs of macronutrient intake in UKBB and CHARGE are available at https://t2d.hugeamp.org/dinspector.html?dataset=Merino2022_DietaryIntake_EU.

GWAS summary statistics of macronutrient intake in only CHARGE are available in dbGaP with accession number phs000930, at https://www.ncbi.nlm.nih.gov/projects/gap/cgi-bin/study.cgi?study_id=phs000930.v10.p1.

GWAS summary statistics of alcohol use (drinks/week) is available at the GWAS catalog (GCST007461) at https://www.ebi.ac.uk/gwas/studies/GCST007461.

## Acknowledgements

This work was supported by grants from the Novo Nordisk Foundation (grant numbers NNF23SA0084103 and NNF18CC0034900).

