## Supplemental tables and figures for "Diet pattern determines circulating FGF21 levels while distinct *FGF21* variants influence diet pattern and FGF21 levels"

**Supplemental table 1.** *CARMA* fine-mapping of the region of chr19:49180000 to 49300000 for FGF21, protein intake, carbohydrate intake and fat intake.

| Trait | Chr | Position | A1 | A0 | EAF | rsID | Consequence | Beta | SE | P-value | PIP |
|---|---|---|---|---|---|---|---|---|---|---|---|
| FGF21 | 19 | 49249663 | G | C | 0.04 | rs28400017 | Intron variant | -0.152 | 0.019 | 2.4E-15 | 0.711 |
| FGF21 | 19 | 49260677 | A | C | 0.54 | rs838131 | Intron variant | 0.123 | 0.008 | 4.7E-58 | 1.000 |
| FGF21 | 19 | 49261038 | G | A | 0.30 | rs1688263 | Intron variant | -0.108 | 0.008 | 7.1E-39 | 0.906 |
| FGF21 | 19 | 49264232 | C | T | 0.09 | rs79390987 | Downstream gene variant | -0.085 | 0.013 | 1.8E-10 | 0.997 |
| FGF21 | 19 | 49273784 | G | A | 0.30 | rs560343 | Intergenic variant | -0.084 | 0.008 | 3.6E-25 | 1.000 |
| Protein | 19 | 49259529 | A | G | 0.44 | rs838133 | Synonymous variant | -0.109 | 0.010 | 2.3E-27 | 1.000 |
| Protein | 19 | 49261368 | T | C | 0.39 | rs739320 | Missense variant | -0.103 | 0.011 | 2.8E-22 | 0.166 |
| Fat | 19 | 49218060 | T | C | 0.48 | rs35866622 | Splice region variant | -0.176 | 0.019 | 6.5E-21 | 0.305 |
| Fat | 19 | 49218111 | T | G | 0.53 | rs33988101 | Synonymous variant | -0.183 | 0.019 | 1.4E-22 | 0.633 |
| Fat | 19 | 49259529 | A | G | 0.44 | rs838133 | Synonymous variant | -0.198 | 0.020 | 1.6E-23 | 1.000 |
| Carbo-hydrate | 19 | 49259529 | A | G | 0.44 | rs838133 | Synonymous variant | 0.210 | 0.025 | 2.4E-17 | 0.994 |

Results of *CARMA* fine-mapping, listing genetic variants that are genome-wide significant ($P<5\times10^{8}$) and with a PIP >0.1.

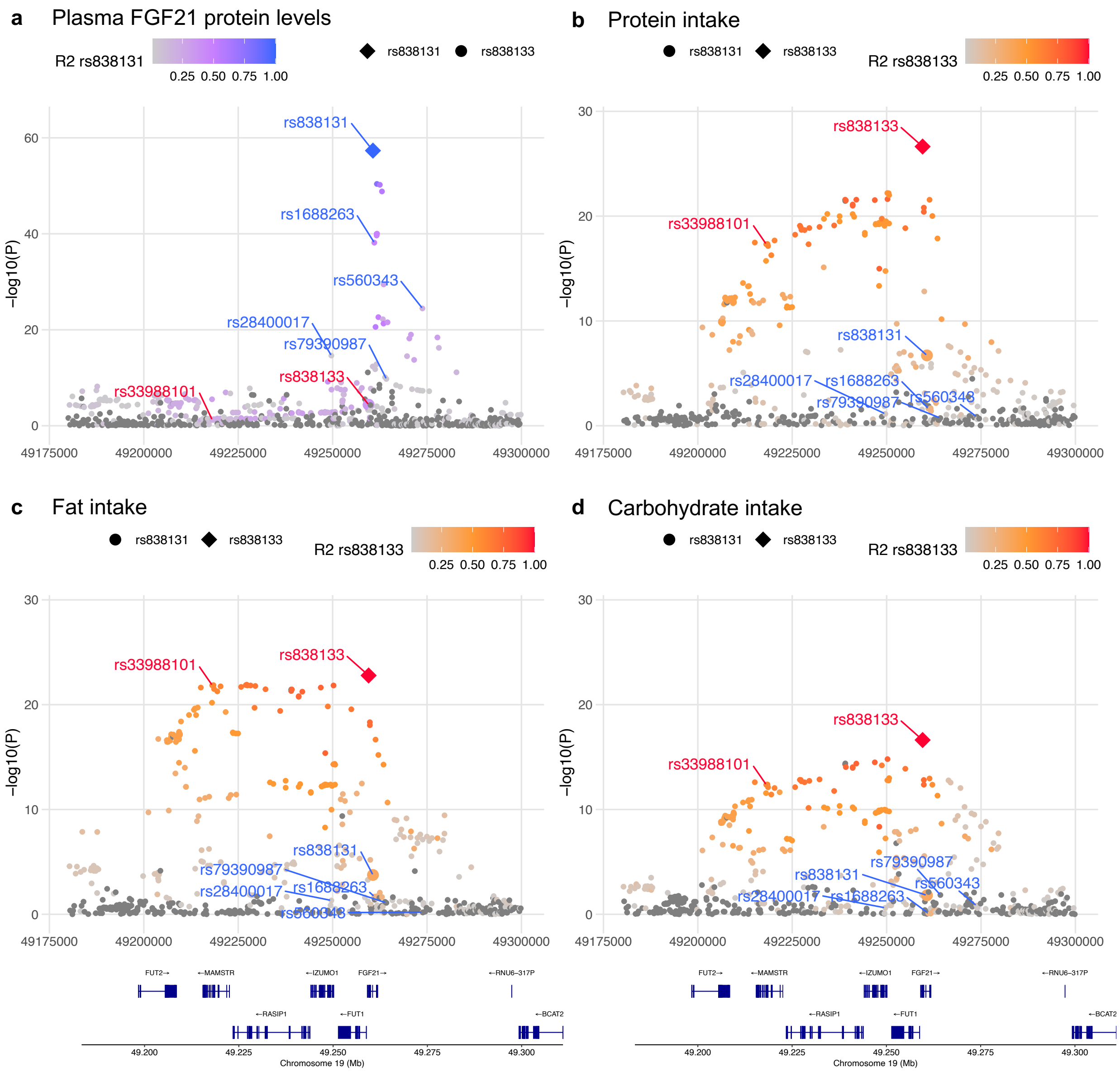


**Supplemental figure 1.** Regional plots of the region 19:49180000 to 19:49300000 for **a**, plasma FGF21 protein levels, **b**, protein intake, **c**, fat intake and **d**, carbohydrate intake, highlighting probable causal variants (PIP>0.5) identified with *CARMA*. Variants in blue are identified as probable causal variants for plasma FGF21 protein levels and variants in red are identified as probable causal variants for macronutrient intake (rs838133 is identified for protein, fat and carbohydrate, while rs33988101 is only identified for fat).

**Supplemental table 2.** Results of colocalization analyses between plasma FGF21 levels and macronutrient intake in the region of chr19:49180000 to 49300000.

| | | **Merino 2022** | | | **Merino 2018** | | |
|---|---|---|---|---|---|---|---|
| | | **CHARGE+UKBB, n=282,271** | | | **CHARGE, n=91,114** | | |
| | | Protein intake | Carbo-hydrate intake | Fat intake | Protein intake | Carbo-hydrate intake | Fat intake |
| | N snps | 609 | 609 | 609 | 368 | 368 | 368 |
| **H0.PP** | No association with either trait | 2.9E-71 | 3.3E-61 | 1.2E-67 | 5.4E-54 | 1.7E-53 | 1.9E-53 |
| **H1.PP** | Association with trait 1, not with trait 2 | 1.1E-20 | 1.3E-10 | 4.5E-17 | 2.1E-03 | 6.6E-03 | 7.6E-03 |
| **H2.PP** | Association with trait 2, not with trait 1 | 2.6E-51 | 2.6E-51 | 2.6E-51 | 2.6E-51 | 2.6E-51 | 2.6E-51 |
| **H3.PP** | Association with trait 1 and trait 2, two independent SNPs | 1.00 | 1.00 | 1.00 | 0.99 | 0.99 | 0.99 |
| **H4.PP** | Association with trait 1 and trait 2, one shared SNP | 1.8E-17 | 4.8E-12 | 9.7E-17 | 6.4E-03 | 2.6E-03 | 2.6E-03 |

Colocalization was analyzed using the *coloc* R-package using the default prior probabilities.

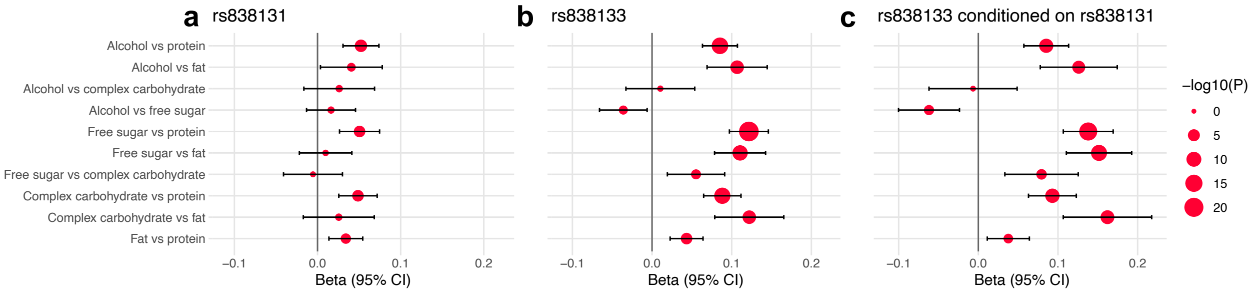


**Supplemental figure 2.** Association between *FGF21* genetic variants and macronutrient intake substitutions. Analyses are adjusted for age, sex, study center, BMI, total energy intake and 10 genetic PCs in addition to macronutrients for substitution modelling (n=146,241-157,508).

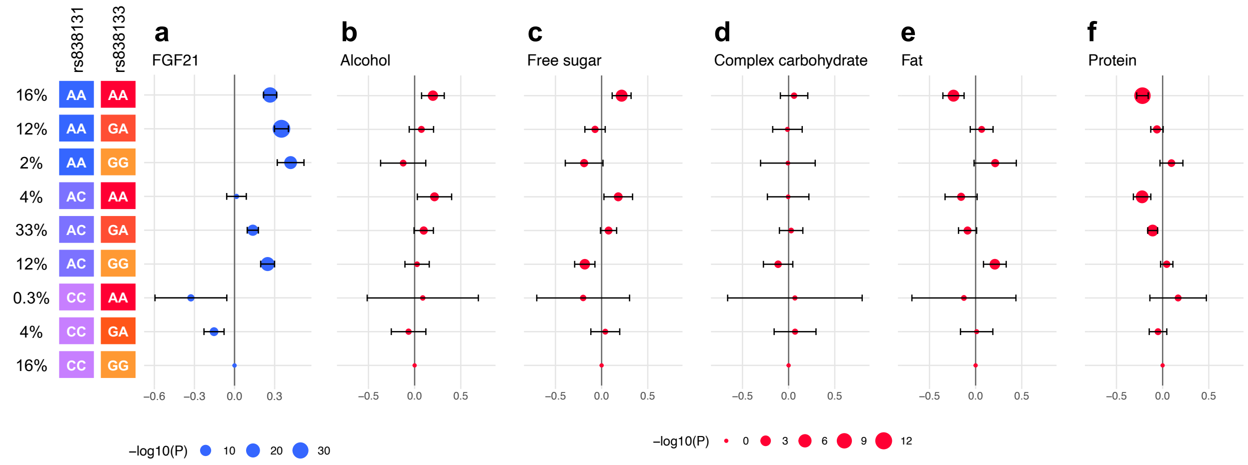


**Supplemental figure 3.** Association between rs838131 and rs838133 genotype group combinations and plasma FGF21 levels and macronutrient intake levels. **a**, Analyses with plasma FGF21 are adjusted for age, sex, study center, fasting time, BMI and 10 genetic PCs (n=36,029). **b-f**, Analyses with macronutrients are adjusted for age, sex, study center, BMI, total energy intake and 10 genetic PCs (n=146,241).

**Supplemental table 3.** Association between macronutrient substitutions and plasma FGF21 protein levels stratified by BMI≥30 or BMI<30.

| **BMI ≥30, n=4585** | **Beta** | **SE** | **P** | **95% CI L** | **95%CI U** |
|---|---|---|---|---|---|
| Alcohol vs protein | 0.047 | 0.006 | 5.3E-16 | 0.059 | 0.036 |
| Alcohol vs fat | 0.016 | 0.004 | 1.5E-05 | 0.023 | 0.009 |
| Alcohol vs complex carbohydrate | 0.022 | 0.003 | 1.4E-13 | 0.028 | 0.016 |
| Alcohol vs free sugar | 0.018 | 0.004 | 1.9E-05 | 0.027 | 0.010 |
| Free sugar vs protein | 0.028 | 0.005 | 2.4E-07 | 0.039 | 0.017 |
| Free sugar vs fat | -0.003 | 0.004 | 4.4E-01 | 0.005 | -0.012 |
| Free sugar vs complex carbohydrate | 0.004 | 0.004 | 2.4E-01 | 0.011 | -0.003 |
| Complex carbohydrate vs protein | 0.024 | 0.006 | 2.9E-05 | 0.035 | 0.013 |
| Complex carbohydrate vs fat | -0.008 | 0.003 | 1.5E-02 | -0.002 | -0.014 |
| Fat vs protein | 0.032 | 0.006 | 2.5E-07 | 0.044 | 0.020 |
| **BMI <30, n=17019** | | | | | |
| Alcohol vs protein | 0.064 | 0.003 | 7.4E-88 | 0.070 | 0.058 |
| Alcohol vs fat | 0.031 | 0.002 | 2.5E-58 | 0.034 | 0.027 |
| Alcohol vs complex carbohydrate | 0.036 | 0.002 | 1.1E-121 | 0.039 | 0.033 |
| Alcohol vs free sugar | 0.022 | 0.002 | 1.2E-23 | 0.026 | 0.017 |
| Free sugar vs protein | 0.041 | 0.003 | 3.2E-40 | 0.047 | 0.035 |
| Free sugar vs fat | 0.007 | 0.002 | 1.5E-03 | 0.012 | 0.003 |
| Free sugar vs complex carbohydrate | 0.014 | 0.002 | 3.3E-13 | 0.018 | 0.010 |
| Complex carbohydrate vs protein | 0.026 | 0.003 | 3.6E-16 | 0.032 | 0.020 |
| Complex carbohydrate vs fat | -0.008 | 0.002 | 8.9E-06 | -0.004 | -0.011 |
| Fat vs protein | 0.033 | 0.003 | 7.8E-23 | 0.040 | 0.027 |

Analyses were adjusted for age, sex, study center, fasting time and total energy intake in addition to macronutrients for substitution modelling.

**Supplemental table 4.** Association between macronutrient substitutions and plasma FGF21 protein levels stratified by fasting time ≥6 h or <6 h.

| **Fasting time ≥6 h, n=2108** | **Beta** | **SE** | **P** | **95% CI L** | **95%CI U** |
|---|---|---|---|---|---|
| Alcohol vs protein | 0.048 | 0.008 | 7.2E-09 | 0.064 | 0.032 |
| Alcohol vs fat | 0.030 | 0.005 | 5.6E-10 | 0.040 | 0.021 |
| Alcohol vs complex carbohydrate | 0.033 | 0.004 | 5.7E-17 | 0.041 | 0.026 |
| Alcohol vs free sugar | 0.024 | 0.006 | 2.7E-05 | 0.034 | 0.013 |
| Free sugar vs protein | 0.023 | 0.008 | 3.8E-03 | 0.038 | 0.007 |
| Free sugar vs fat | 0.005 | 0.006 | 3.9E-01 | 0.017 | -0.006 |
| Free sugar vs complex carbohydrate | 0.010 | 0.005 | 4.7E-02 | 0.019 | 0.000 |
| Complex carbohydrate vs protein | 0.012 | 0.008 | 1.3E-01 | 0.029 | -0.004 |
| Complex carbohydrate vs fat | -0.005 | 0.005 | 2.4E-01 | 0.004 | -0.014 |
| Fat vs protein | 0.018 | 0.009 | 4.3E-02 | 0.035 | 0.001 |
| **Fasting time <6 h, n=17019** | | | | | |
| Alcohol vs protein | 0.067 | 0.003 | 3.3E-116 | 0.073 | 0.061 |
| Alcohol vs fat | 0.027 | 0.002 | 1.8E-52 | 0.030 | 0.023 |
| Alcohol vs complex carbohydrate | 0.032 | 0.001 | 1.0E-114 | 0.035 | 0.029 |
| Alcohol vs free sugar | 0.021 | 0.002 | 1.5E-25 | 0.025 | 0.017 |
| Free sugar vs protein | 0.045 | 0.003 | 1.7E-58 | 0.050 | 0.039 |
| Free sugar vs fat | 0.004 | 0.002 | 4.3E-02 | 0.008 | 0.000 |
| Free sugar vs complex carbohydrate | 0.011 | 0.002 | 2.4E-10 | 0.015 | 0.008 |
| Complex carbohydrate vs protein | 0.033 | 0.003 | 2.4E-30 | 0.039 | 0.027 |
| Complex carbohydrate vs fat | -0.008 | 0.002 | 1.2E-06 | -0.005 | -0.011 |
| Fat vs protein | 0.041 | 0.003 | 2.4E-39 | 0.047 | 0.035 |

Analyses were adjusted for age, sex, study center, BMI and total energy intake in addition to macronutrients for substitution modelling.

**Supplemental table 5.** Association between macronutrient substitutions and plasma FGF21 protein levels adjusting for *FGF21* genetic variation.

| **Unadjusted** | **Beta** | **SE** | **P** | **95% CI L** | **95%CI U** |
|---|---|---|---|---|---|
| Alcohol vs protein | 0.066 | 0.003 | 3.9E-87 | 0.060 | 0.073 |
| Alcohol vs fat | 0.027 | 0.002 | 3.0E-43 | 0.023 | 0.030 |
| Alcohol vs complex carbohydrate | 0.035 | 0.002 | 5.2E-99 | 0.032 | 0.039 |
| Alcohol vs free sugar | 0.023 | 0.002 | 6.9E-22 | 0.019 | 0.028 |
| Free sugar vs protein | 0.040 | 0.003 | 2.4E-41 | 0.034 | 0.046 |
| Free sugar vs fat | 0.003 | 0.002 | 0.176 | -0.001 | 0.008 |
| Free sugar vs complex carbohydrate | 0.011 | 0.002 | 1.8E-08 | 0.007 | 0.015 |
| Complex carbohydrate vs protein | 0.029 | 0.003 | 3.9E-20 | 0.023 | 0.035 |
| Complex carbohydrate vs fat | -0.008 | 0.002 | 7.8E-07 | -0.012 | -0.005 |
| Fat vs protein | 0.039 | 0.004 | 6.3E-29 | 0.033 | 0.046 |
| **Adjusted for rs838131** | | | | | |
| Alcohol vs protein | 0.065 | 0.003 | 6.0E-86 | 0.059 | 0.072 |
| Alcohol vs fat | 0.026 | 0.002 | 3.6E-41 | 0.022 | 0.030 |
| Alcohol vs complex carbohydrate | 0.035 | 0.002 | 1.9E-97 | 0.032 | 0.038 |
| Alcohol vs free sugar | 0.023 | 0.002 | 7.9E-22 | 0.019 | 0.028 |
| Free sugar vs protein | 0.039 | 0.003 | 1.5E-40 | 0.034 | 0.045 |
| Free sugar vs fat | 0.002 | 0.002 | 0.271 | -0.002 | 0.007 |
| Free sugar vs complex carbohydrate | 0.011 | 0.002 | 3.6E-08 | 0.007 | 0.015 |
| Complex carbohydrate vs protein | 0.028 | 0.003 | 6.4E-20 | 0.022 | 0.034 |
| Complex carbohydrate vs fat | -0.009 | 0.002 | 2.7E-07 | -0.012 | -0.005 |
| Fat vs protein | 0.039 | 0.004 | 3.3E-29 | 0.033 | 0.046 |
| **Adjusted for rs838133** | | | | | |
| Alcohol vs protein | 0.066 | 0.003 | 1.1E-85 | 0.059 | 0.072 |
| Alcohol vs fat | 0.026 | 0.002 | 1.6E-41 | 0.022 | 0.030 |
| Alcohol vs complex carbohydrate | 0.035 | 0.002 | 2.9E-97 | 0.032 | 0.038 |
| Alcohol vs free sugar | 0.023 | 0.002 | 2.3E-21 | 0.018 | 0.028 |
| Free sugar vs protein | 0.040 | 0.003 | 8.3E-41 | 0.034 | 0.046 |
| Free sugar vs fat | 0.003 | 0.002 | 0.209 | -0.002 | 0.007 |
| Free sugar vs complex carbohydrate | 0.011 | 0.002 | 2.0E-08 | 0.007 | 0.015 |
| Complex carbohydrate vs protein | 0.028 | 0.003 | 7.8E-20 | 0.022 | 0.035 |
| Complex carbohydrate vs fat | -0.008 | 0.002 | 4.2E-07 | -0.012 | -0.005 |
| Fat vs protein | 0.039 | 0.004 | 6.5E-29 | 0.033 | 0.046 |
| **Adjusted for rs838131 and rs838133** | | | | | |
| Alcohol vs protein | 0.066 | 0.003 | 3.4E-88 | 0.060 | 0.073 |
| Alcohol vs fat | 0.026 | 0.002 | 8.9E-42 | 0.022 | 0.030 |
| Alcohol vs complex carbohydrate | 0.035 | 0.002 | 3.4E-97 | 0.032 | 0.038 |
| Alcohol vs free sugar | 0.023 | 0.002 | 2.3E-21 | 0.018 | 0.028 |
| Free sugar vs protein | 0.040 | 0.003 | 1.2E-42 | 0.035 | 0.046 |
| Free sugar vs fat | 0.003 | 0.002 | 0.196 | -0.002 | 0.007 |
| Free sugar vs complex carbohydrate | 0.011 | 0.002 | 2.0E-08 | 0.007 | 0.015 |

| | | | | | |
|---|---|---|---|---|---|
| Complex carbohydrate vs protein | 0.029 | 0.003 | 4.5E-21 | 0.023 | 0.035 |
| Complex carbohydrate vs fat | -0.008 | 0.002 | 5.6E-07 | -0.012 | -0.005 |
| Fat vs protein | 0.040 | 0.004 | 3.4E-30 | 0.033 | 0.047 |
| **Adjusted for weigthed SNP-score of 7 pQTLs** | | | | | |
| Alcohol vs protein | 0.066 | 0.003 | 3.4E-85 | 0.060 | 0.073 |
| Alcohol vs fat | 0.026 | 0.002 | 4.6E-40 | 0.022 | 0.030 |
| Alcohol vs complex carbohydrate | 0.035 | 0.002 | 1.2E-93 | 0.032 | 0.038 |
| Alcohol vs free sugar | 0.024 | 0.002 | 3.5E-22 | 0.019 | 0.029 |
| Free sugar vs protein | 0.040 | 0.003 | 7.4E-40 | 0.034 | 0.046 |
| Free sugar vs fat | 0.002 | 0.002 | 0.376 | -0.002 | 0.007 |
| Free sugar vs complex carbohydrate | 0.010 | 0.002 | 2.6E-07 | 0.006 | 0.014 |
| Complex carbohydrate vs protein | 0.029 | 0.003 | 2.1E-20 | 0.023 | 0.035 |
| Complex carbohydrate vs fat | -0.008 | 0.002 | 6.5E-07 | -0.012 | -0.005 |
| Fat vs protein | 0.040 | 0.004 | 2.5E-29 | 0.033 | 0.047 |

Analyses were adjusted for age, sex, study center, fasting time, BMI, total energy intake and 10 genetics PCs in addition to macronutrients for substitution modelling and the indicated genetic variants.

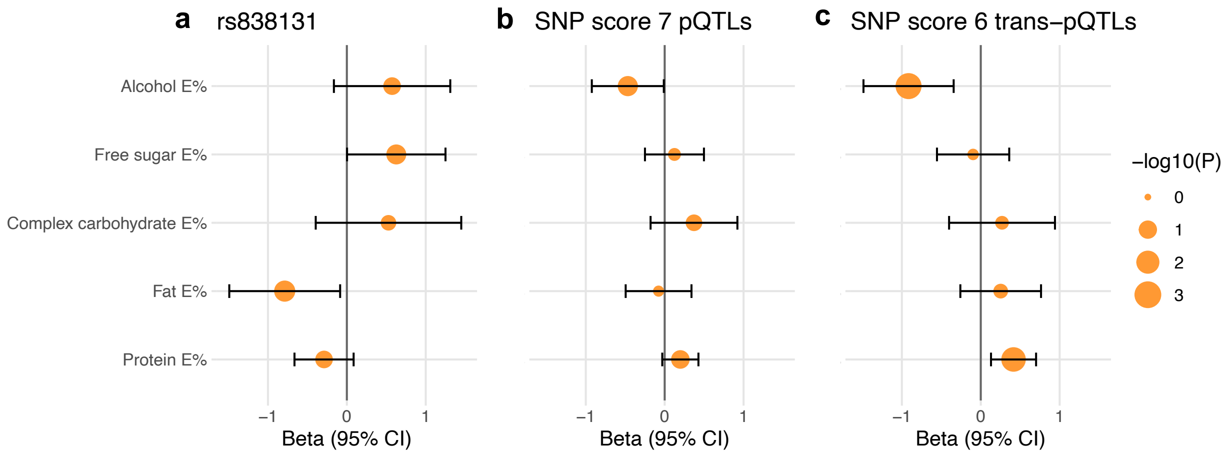


**Supplemental figure 4.** One-sample MR analyzes estimating potentially causal effects of plasma FGF21 on macronutrient intake using different IVs: **a**, the FGF21 cis-pQTL (rs838131), **b**, all 7 FGF21 pQTLS composed into a weighted score, and **c**, only the 6 trans-pQTLs (excluding rs838131) composed into a weighted score. Analyses were adjusted for age, sex, study center, fasting time, BMI, total energy intake and 10 genetic PCs (n=15,512-16,400).

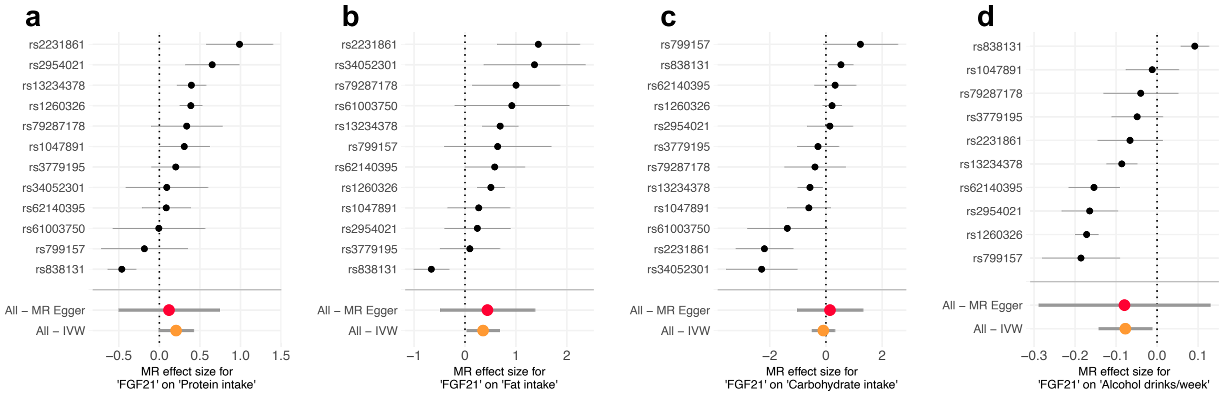


**Supplemental figure 5.** Forest plot of single IV estimates in two-sample MR estimating the potentially causal effect of plasma FGF21 on **a**, protein intake, **b**, fat intake, **c**, carbohydrate intake, and **d**, alcohol intake.

**Supplemental table 6.** Two-sample MR heterogeneity when including or excluding the rs838131 variant as IV.

| | All IVs | | | | Excluding rs838131 | | | |
|---|---|---|---|---|---|---|---|---|
| | Q | df | P | $i^2$ | Q | df | P | $i^2$ |
| Protein | | | | | | | | |
| IVW | 93.1 | 11 | 4.2E-15 | 88.2% | 23.4 | 10 | 9.2E-03 | 57.4% |
| Egger | 92.3 | 10 | 1.9E-15 | 89.2% | 23.4 | 9 | 5.5E-03 | 61.5% |
| Fat | | | | | | | | |
| IVW | 53.6 | 11 | 1.4E-07 | 79.5% | 13.3 | 10 | 2.1E-01 | 24.7% |
| Egger | 53.4 | 10 | 6.3E-08 | 81.3% | 11.4 | 9 | 2.5E-01 | 21.2% |
| Carbohydrate | | | | | | | | |
| IVW | 54.3 | 11 | 6.3E-08 | 79.8% | 44.2 | 10 | 2.7E-06 | 77.4% |
| Egger | 53.4 | 10 | 1.0E-07 | 81.3% | 44.5 | 9 | 1.3E-06 | 79.8% |
| Alcohol | | | | | | | | |
| IVW | 160.1 | 8 | 7.1E-30 | 95.0% | 41.3 | 8 | 2.4E-06 | 80.6% |
| Egger | 160.1 | 9 | 1.5E-30 | 94.4% | 38.6 | 7 | 1.8E-06 | 81.8% |

Heterogeneity was tested with the *mr_heterogeneity* function for both the IVW model and the MR Egger model. $I^2$ was calculated as 100*(Q-df)/Q.
